# PreTA: A network meta-analysis ranking metric measuring the probability of being preferable than the average treatment

**DOI:** 10.1101/2020.04.18.20070615

**Authors:** Adriani Nikolakopoulou, Dimitris Mavridis, Virginia Chiocchia, Theodoros Papakonstantinou, Toshi A Furukawa, Georgia Salanti

## Abstract

**Background:** Network meta-analysis (NMA) produces complex outputs as many comparisons between interventions are of interest and a treatment ranking is often included in the aims of the evidence synthesis. The estimated relative treatment effects are usually displayed in a forest plot or in a league table and several ranking metrics are calculated and presented, such as the median and mean treatment ranks.

**Methods:** We estimate relative treatment effects of each competing treatment against a fictional ‘average’ treatment using the ‘deviation from the means’ coding that has been used to parametrize categorical covariates in regression models. Based on this alternative parametrization of the NMA model, we present a new ranking metric (PreTA: Preferable Than Average) interpreted as the probability that a treatment is better than a fictional treatment of average performance.

**Results:** We compare PreTA with existing probabilistic ranking metrics in 232 networks of interventions. We use two networks of interventions, a network of 18 antidepressants for acute depression and a network of four interventions for heavy menstrual bleeding, to illustrate the methodology. The agreement between PreTA and existing ranking metrics depends on the precision with which relative effects are estimated.

**Conclusions:** PreTA is a viable alternative to existing ranking metrics which can be interpreted as the probability of being better than the ‘average’ treatment. It enriches the decision-making arsenal with a ranking metric which is interpreted as a probability and considers the entire ranking distributions of the involved treatments.

## 1 Introduction

Results from network meta-analysis (NMA) are often used to inform health-care decision making and their presentation in a coherent and understandable way is of critical importance (1,2). The main output of NMA is a set of relative effects between all treatments, which has been produced by combining direct and indirect evidence in a network of trials comparing different treatments (3,4). A very informative way of presenting the NMA relative treatment effects is in a league table, where the names of the treatments are presented in the diagonal and each cell contains the relative treatment effect (5). Such a table allows for the simultaneous presentation of two outcomes, or of the results from pairwise and network meta-analysis, below and above the diagonal.

Although the set of relative effects contains all the information produced from NMA, a treatment hierarchy is often of interest to decision makers and end users. To this aim, alongside treatment effect estimates, several ranking metrics have been proposed to present NMA results. Ranking probabilities of each treatment being at each possible rank are calculated using simulation or resampling techniques either in a Bayesian or in a frequentist framework. Other ranking metrics include the surface under the cumulative ranking curve (SUCRA), that averages across all ranking probabilities for each treatment, and its frequentist analogue, P-score, which is calculated analytically (6,7). SUCRA and P-score can be interpreted as the mean extent of certainty that a treatment is better than all the other treatments. As authors of (6) point out, however, *“it is impossible to tell what constitutes a modest or large difference in SUCRA between two treatments, either statistically or clinically”*. An alternative way to produce a treatment hierarchy is to simply rank treatments according to the relative effects versus placebo, or another reference treatment. However, this hierarchy either does not take into account uncertainty (by considering only point estimates) or depends a lot on the uncertainty around the reference treatment.

In this paper, we develop a probabilistic ranking metric that naturally incorporates uncertainty and is a viable alternative to existing ranking metrics. We re-parametrize the NMA model to derive treatment effects against a fictional treatment of average performance using the deviation of means coding that has been used to parametrize categorical covariates in regression models (8). Then, we use the derived treatment effects to compute the probability of each treatment being better than the ‘average’ treatment. This ranking metric aids the interpretation of NMA results in classifying treatments as superior, equivalent and inferior to an imaginary ‘average’ treatment.

## 2 Reparametrization of the NMA model

### 2.1 Deviation from means coding in regression models

We start with a short description of the deviation from means coding in regression models as described by Hosmer and Lemeshow (8). This is an alternative parametrization to the most common ‘reference cell coding’ in order to avoid the use of a reference level. According to the reference cell coding, a categorical independent variable with *C* categories is expressed through *C* − 1 dummy/indicator variables.

Consider, for example, that we aim to estimate the effect of a covariate with four groups on the probability of an event. We fit a logistic regression model

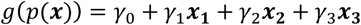

where ***x*** = (***x***_1_, ***x***_2_, ***x***_3_)′ are the dummy variables for the covariate and *g*(*p*(***x***)) is the logit link function *g*(*p*(***x***)) = *logit*(*p*(***x***)) = *log*(*p*(***x***)/(1 − *p*(***x***))) with *p*(***x***) indicating the probability of event.

According to the reference cell coding, the indicator variables are parametrized as shown in Table 1 and result into estimating logarithms of the relative odds ratios (logOR) between the categories represented by the values 0 and 1 in these indicator variables.

**Table 1.** Illustration of construction of dummy variables for modelling a categorical variable with four groups in regression using reference cell coding and deviation from means coding.

| Reference cell coding |  |  |  | Deviation from means coding |  |  |  |
| --- | --- | --- | --- | --- | --- | --- | --- |
| Dummy variables |  |  |  | Dummy variables |  |  |  |
| Covariate | $x_1$ | $x_2$ | $x_3$ | Covariate | $x_1^*$ | $x_2^*$ | $x_3^*$ |
| Group 1 | 0 | 0 | 0 | Group 1 | -1 | -1 | -1 |
| Group 2 | 1 | 0 | 0 | Average* | 0 | 0 | 0 |
| Group 3 | 0 | 1 | 0 | Group 2 | 1 | 0 | 0 |
| Group 4 | 0 | 0 | 1 | Group 3 | 0 | 1 | 0 |
|  |  |  |  | Group 4 | 0 | 0 | 1 |

According to the alternative deviation from means coding, the indicator variables express effects as deviations between each category mean (here the logit of the outcome in that category) from the overall (grand) mean (here the average logit outcome over all categories). We re-write the model as

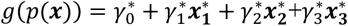

where the indicator variables 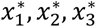 are defined as shown in Table 1. The model results in estimating the coefficients 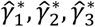, interpreted as the relative effects among groups versus the average effect across all groups. Note that the exponential of the coefficients 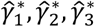 are not odds ratios because in the denominator is the average odds that includes the odds of the numerator.

For example, the logit of group 2 versus average logit over all groups is derived as

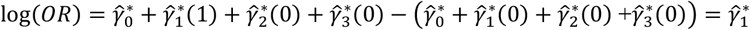

For further information and examples on the deviation from means coding, see (8).

### 2.2 Notation for the NMA model

In this section, we introduce some general notation for the NMA model. Let the entire evidence base consist of *i* = 1, …, *n* studies forming a set of treatments, denoted as *k* = 1, …, *K*. The number of treatments in study *i* is denoted as *K*_*i*_. Index *j* denotes a treatment contrast. A core assumption in NMA is that of transitivity, which implies that in a network of *K* treatments, and subsequently 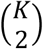 possible relative treatment effects, only *K* − 1 need to be estimated and the rest are derived as linear combinations of those (9,10). The target parameter is therefore a vector ***μ*** of *K* − 1 relative treatment effects *μ*_**2**_, *μ*_3_, … *μ*_*K*_, called the vector of basic parameters (11,12).

With arm-level data we can model arm level parameters, for example the event probability for a binary outcome, in study *i* and treatment arm *k* denoted as *y*_*ik*_(13). A link function *g*(*y*_*ik*_) maps the parameters of interest onto a scale ranging from minus to plus infinity and *u*_*i*_ are the trial-specific baselines. For an overview of commonly used link functions in meta-analysis see (14). All arm-level parameters *y*_*ik*_ across studies are collected in a vector ***y***^***a***^ of length 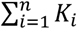, where superscript *a* stands for ‘arm-level’.

With contrast-level data we model trial specific summaries, for example logOR, log risk ratio, mean difference or standardized mean difference (13). Let *y*_*ij*_ be the observed effect size for treatment contrast *j* in study *i*. The vector of the estimated contrasts across all studies is denoted as ***y***^***c***^ and is of length 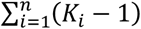. The superscript ***c*** indicates the fact that ‘contrast-level’ data are modeled.

We will first describe the arm-level and then the contrast-level NMA models using reference cell coding and the equivalent alternative deviation from the means parametrization, which allows estimation of all treatments versus a fictional treatment of average performance. We will exemplify the models using a hypothetical network of three treatments, A, B and C examined in four studies, one comparing A and B, one comparing A and C, one comparing B and C and one three-arm study comparing treatments A, B and C. The target vector of basic parameters is usually taken to include the relative effects of all treatments versus an arbitrary reference, here treatment A, and hence is 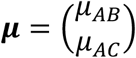. The transitivity assumption implies consistency between relative treatment effects; in particular, it holds that

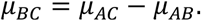

### 2.3 NMA with arm-level data

#### 2.3.1 Reference cell coding

The model for study 1, comparing treatments A and B is shown in Table 2; *δ*_1,*AB*_ denotes the random effect of study 1 for the comparison AB and *τ*^2^ denotes heterogeneity. It is customary to assume that heterogeneity is common across comparisons. The model is straightforwardly generalized for the other three studies (Table 2).

**Table 2.** Arm-level and contrast-level NMA models using reference cell coding and deviation from means coding for a fictional network of three treatments examined in four studies.

| Study number,<br>treatments<br>compared | Arm-based NMA |  | Contrast-based NMA |  |
| --- | --- | --- | --- | --- |
|  | Reference cell coding | Deviation from means coding | Reference cell coding | Deviation from means coding |
| Study 1, AB | $g(y_{1A}) = u_1$<br>$g(y_{1B}) = u_1 + \mu_{AB} + \delta_{1,AB}$<br>$\delta_{1,AB} \sim N(0, \tau^2)$ | $g(y_{1A}) = u_1 - b_B - b_C$<br>$g(y_{1B}) = u_1 + b_B + \delta_{1,AB}$<br>$\delta_{1,AB} \sim N(0, \tau^2)$ | $y_{1,AB} = \mu_{AB} + \varepsilon_{1,AB} + \delta_{1,AB}$<br>$\varepsilon_{1,AB} \sim N(0, s_{1,AB}^2)$<br>$\delta_{1,AB} \sim N(0, \tau^2)$ | $y_{1,AB} = 2b_B + b_C + \varepsilon_{1,AB} + \delta_{1,AB}$<br>$\varepsilon_{1,AB} \sim N(0, s_{1,AB}^2)$<br>$\delta_{1,AB} \sim N(0, \tau^2)$ |
| Study 2, AC | $g(y_{2A}) = u_2$<br>$g(y_{2C}) = u_2 + \mu_{AC} + \delta_{2,AC}$<br>$\delta_{2,AC} \sim N(0, \tau^2)$ | $g(y_{2A}) = u_2 - b_B - b_C$<br>$g(y_{2C}) = u_2 + b_C + \delta_{2,AC}$<br>$\delta_{2,AC} \sim N(0, \tau^2)$ | $y_{2,AC} = \mu_{AC} + \varepsilon_{2,AC} + \delta_{2,AC}$<br>$\varepsilon_{2,AC} \sim N(0, s_{2,AC}^2)$<br>$\delta_{2,AC} \sim N(0, \tau^2)$ | $y_{2,AC} = b_B + 2b_C + \varepsilon_{2,AC} + \delta_{2,AC}$<br>$\varepsilon_{2,AC} \sim N(0, s_{2,AC}^2)$<br>$\delta_{2,AC} \sim N(0, \tau^2)$ |
| Study 3, BC | $g(y_{3B}) = u_3$<br>$g(y_{3C}) = u_3 - \mu_{AB} + \mu_{AC} + \delta_{3,BC}$<br>$\delta_{3,BC} \sim N(0, \tau^2)$ | $g(y_{3B}) = u_3 + b_B$<br>$g(y_{3C}) = u_3 + b_C + \delta_{3,BC}$<br>$\delta_{3,BC} \sim N(0, \tau^2)$ | $y_{3,BC} = -\mu_{AB} + \mu_{AC} + \varepsilon_{3,BC} + \delta_{3,BC}$<br>$\varepsilon_{3,BC} \sim N(0, s_{3,BC}^2)$<br>$\delta_{3,BC} \sim N(0, \tau^2)$ | $y_{3,BC} = -b_B + b_C + \varepsilon_{3,BC} + \delta_{3,BC}$<br>$\varepsilon_{3,BC} \sim N(0, s_{3,BC}^2)$<br>$\delta_{3,BC} \sim N(0, \tau^2)$ |
| Study 4, ABC | $g(y_{4A}) = u_4$<br>$g(y_{4B}) = u_4 + \mu_{AB} + \delta_{4,AB}$<br>$g(y_{4C}) = u_4 + \mu_{AC} + \delta_{4,AC}$<br>$\delta_{4,AB} \sim N(0, \tau^2)$<br>$\delta_{4,AC} \sim N(0, \tau^2)$ | $g(y_{4A}) = u_4 - b_B - b_C$<br>$g(y_{4B}) = u_4 + b_B + \delta_{4,AB}$<br>$g(y_{4C}) = u_4 + b_C + \delta_{4,AC}$<br>$\delta_{4,AB} \sim N(0, \tau^2)$<br>$\delta_{4,AC} \sim N(0, \tau^2)$ | $y_{4,AB} = \mu_{AB} + \varepsilon_{4,AB} + \delta_{4,AB}$<br>$y_{4,AC} = \mu_{AC} + \varepsilon_{4,AC} + \delta_{4,AC}$<br>$\varepsilon_{4,AB} \sim N(0, s_{4,AB}^2)$<br>$\delta_{4,AB} \sim N(0, \tau^2)$<br>$\varepsilon_{4,AC} \sim N(0, s_{4,AC}^2)$<br>$\delta_{4,AC} \sim N(0, \tau^2)$ | $y_{4,AB} = 2b_B + b_C + \varepsilon_{4,AB} + \delta_{4,AB}$<br>$y_{4,AC} = b_B + 2b_C + \varepsilon_{4,AC} + \delta_{4,AC}$<br>$\varepsilon_{4,AB} \sim N(0, s_{4,AB}^2)$<br>$\delta_{4,AB} \sim N(0, \tau^2)$<br>$\varepsilon_{4,AC} \sim N(0, s_{4,AC}^2)$<br>$\delta_{4,AC} \sim N(0, \tau^2)$ |

In its general form, the NMA model using arm-based analysis can be written as

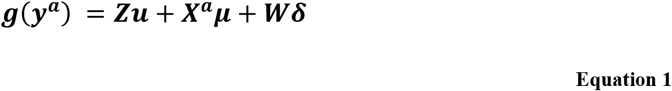

where ***u*** is the vector of baselines *u*_*i*_ of length *n*, which can be assumed to be either fixed and unrelated to each other, or exchangeable drawn from a normal distribution (15). We assume fixed and unrelated baseline effects for the remainder of this paper. Vector ***δ***; includes the study random effects *δ*_*i,j*_ and follows the multivariate normal distribution

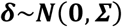

Matrix ***Σ*** is a block-diagonal between-study variance-covariance matrix of dimensions 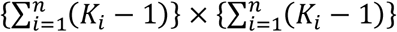. The matrices ***Z, X***^***a***^, ***W*** are design matrices linking the vector of baselines, basic parameters and random effects respectively with ***g***(***y***^***a***^). The construction of these design matrices depends on the modeled arm-level parameters *y*_*ik*_ and is exemplified in the following example.

For the example of Table 2, Equation 1 takes the form

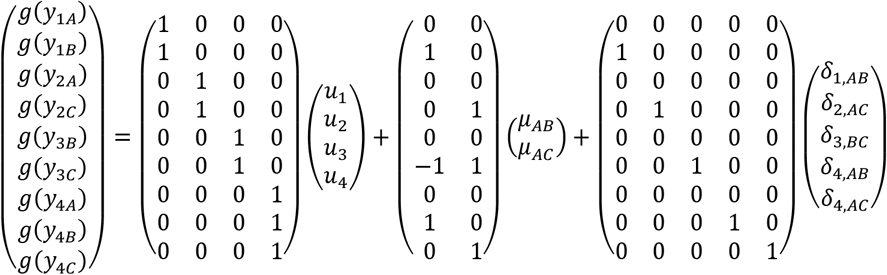

With

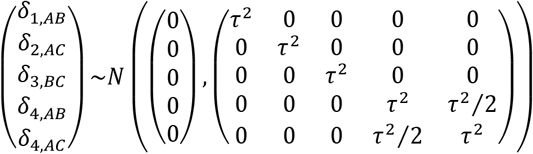

Matrix ***X***^***a***^ indicates which elements of ***μ*** are estimated by each *g*(*y*_*ik*_). It contains one row per study arm and one column per basic parameter. The first row corresponds to treatment arm A of the first study taking the value 0 both for *μ*_*AB*_ and *μ*_*AC*_. The second row indicates that *μ*_*AB*_ is estimated in treatment arm B of the first study. Similarly, the construction of the next rows of ***X***^***a***^, as well as that of ***Z*** and ***W***, is implied by the arm-level data included in each study and the subsequent elements of ***μ*** to be estimated (Table 2).

#### 2.3.2 Deviation from means coding

The above model in Equation 1 can be modified using the deviation from means coding (8). The model will be parametrized in such a way to estimate the effects of each treatment versus the ‘average’ treatment. The target parameter of this model is a vector ***b*** that includes *K* − 1 parameters *b*_*k*_ with *k* = 2, …, *K* which are the effects of treatment *k* versus the average effect over all treatments. One of the treatments – here treatment 1 – is arbitrarily chosen to be excluded for identifiability. Results do not depend on the choice of this ‘reference’ treatment.

For the deviation from means coding, the model will be

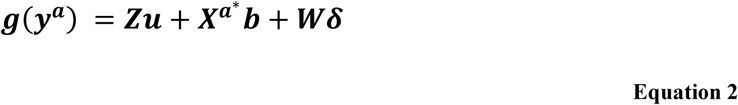

with 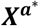 denoting the modified design matrix. The matrices ***Z*** and ***W*** remain unchanged. The new design matrix 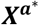 will take values −1 for the arbitrarily chosen treatment that is not included in vector ***b***; all other entries in the matrix are as in ***X***^***a***^.

Consider the example of Table 1 and the first two rows of the ***X***^***a***^ matrix, 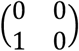, corresponding to the first study. According to the deviation from means coding as illustrated in Table 1, we chose a treatment (here treatment A) for which 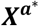 will take −1 for both dummy variables (both columns of the design matrix) and the group corresponding to treatment B takes 1 and 0 for the two columns of the design matrix, as in ***X***^***a***^. Thus, the respective part of the new design matrix will be 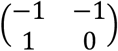. The model for study 1 with the alternative parametrization is

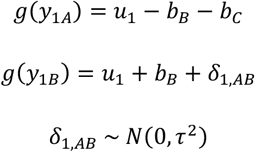

where the parameters *b*_*B*_ and *b*_*C*_ denote the effects of B versus average treatment and C versus average treatment respectively. The effect of A versus the average treatment is −*b*_*B*_ − *b*_*C*_ and the relative effect of B versus A for the study 1 is derived as

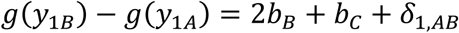

The models for all studies are given in Table 2 and the full model is written as

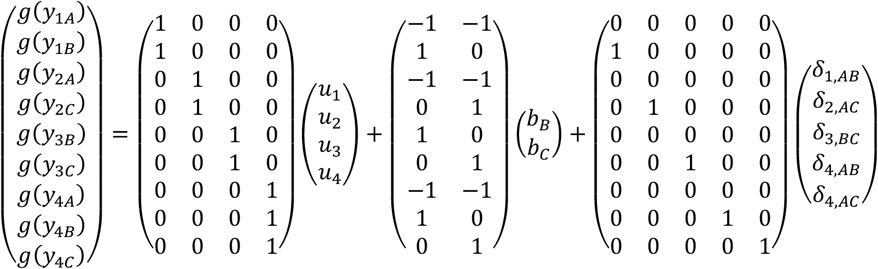

Note that the reparametrization described using the deviation from the means coding should not be confused with different parametrizations of the NMA model to produce relative treatment effects of all treatments versus each other. We present in the Additional file 1 an example of different parametrizations for specifying the means using reference cell coding and deviation from means coding using arm-level data.

### 2.3 NMA with contrast-level data

#### 2.4.1 Reference cell coding

In the contrast-level NMA, data from *K*_*i*_ − 1 contrasts for each study are modeled. The model for study *i* and treatment contrast *j* is written as

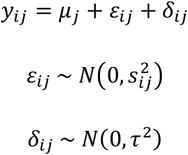

with *ε*_*ij*_ being the random error for study *i* and treatment contrast *j* where 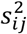 is the sample variance of *y*_*ij*_. The random effect *δ*_*ij*_ is defined as in the NMA with arm-level data. For example, for the first study the model is

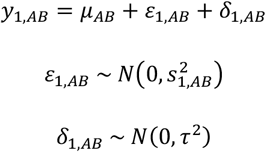

and, similarly, for the other studies the models are given in Table 2.

The contrast-based NMA model in its general form is then written as

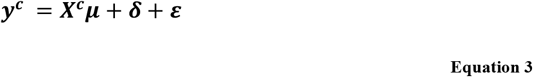

with the vector of random effects ***δ*** having the distribution given in the arm-level NMA model and the vector of random errors being distributed as

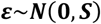

where ***S*** is the block-diagonal within-study variance-covariance matrix of the same dimensions as **∑**. The design matrix ***X***^***c***^ has dimensions 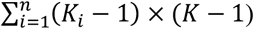. The entries in each row describe the relationship between the vector of basic parameters ***μ*** and the vector of observed contrast-level data ***y***^***c***^.

For example, in the illustrative network of three treatments and four studies, the full model is written as

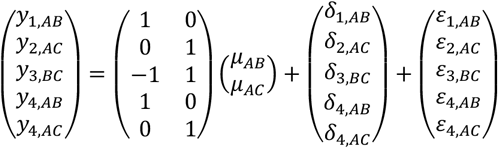

The first row of the ***X***^***c***^ matrix indicates that the first two-arm study estimates *μ*_*AB*_. Note that the arm-level model using reference cell coding for study 1 implies that

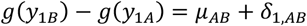

and, consequently, the first row of the ***X***^***c***^ matrix results as the subtraction of the second minus the first row of ***X***^***a***^.

#### 2.4.2 Deviation from means coding

The reparametrized model will differ from that presented in Equation 3 in two ways; the target parameter to be estimated, which again are the relative effects ***b*** against an ‘average’ treatment, and the design matrix 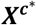. The matrix 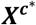 can be easily obtained from ***X***^***a***∗^ by subtracting its rows within each study contrast. In its general form, the model is

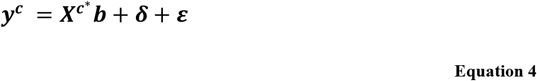

Consider in our example the part of ***X***^***a***\*^ corresponding to study 1,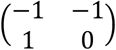, then the row of 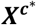 corresponding to that first study will be (2 1), which is the subtraction of the two rows. This is also evident considering that

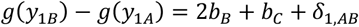

according to the arm-based model using the deviation from means coding.

The models for studies 1 to 4 are given in Table 2 and can be written as

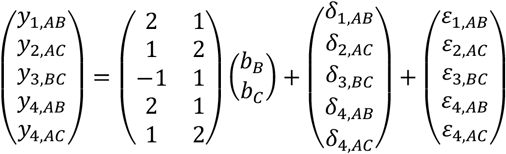

The estimation of ***b*** in the contrast-based NMA model using deviation from means coding (Equation 4) is

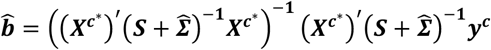

with variance-covariance matrix

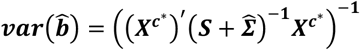

Vector 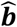 includes the estimation of the *K* − 1 parameters *b*_*k*_ for *k* = 2, …, *K*. The estimation of the effect of treatment *k* = 1, which was chosen to be excluded for identifiability, versus the average effect is given as

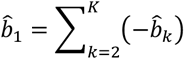

with variance 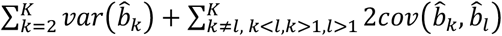. Note that results do not depend on the choice of reference treatment.

Network estimates 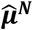 can be derived as linear combinations of 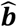

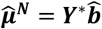

with variance-covariance matrix

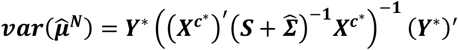

and are equivalent to the network estimates derived using reference cell coding. Matrix ***Y***^*^ of dimensions 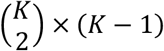 is constructed similarly to 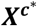 and connects 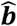 with network estimates 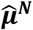. We can use several methods for estimating ***Σ*** such as likelihood-based methods and an extension of the DerSimonian and Laird method (11,16). For the worked example, it holds that

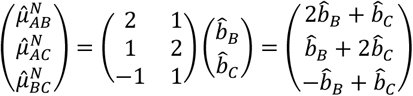

The contrast-level NMA model can be written as a two-stage model, as first described in (11,17,18), where results of separate pairwise meta-analyses are used instead of ***y***^***c***^ in the model described in Equation 3. Constructing the respective design matrix follows the logic of constructing ***X***^***c***^ and its modification to parametrize the model using the deviation from means coding is straightforward.

## 3 PreTA: Probability of a treatment being preferable than the average treatment

Applying the deviation from means coding in NMA models results into the derivation of the effects of each treatment against a fictional treatment of ‘average’ performance. In this section we use the *K* estimated parameters 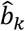 to compute the probability of each treatment being better than the average treatment. To do so, we follow similar steps as those followed by Rücker and Schwarzer who derived the frequentist analogue of SUCRA, P-score (7).

Intermediate to the calculation of P-scores is the derivation of the probability that treatment *k* is better than treatment *l*, calculated as

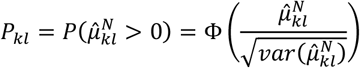

assuming that higher values represent a better outcome. Accordingly, the probability that treatment *k* is better than the fictional treatment of average performance (PreTA) can be derived as

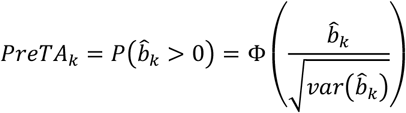

The range of values for *PreTA*_*k*_ is (0.5, 1) if 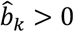, and (0, 0.5) if 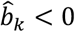. As it is the case with P-scores, the mean of *beta*_*k*_ across all treatments is 0.5. Alternatively, the z-score 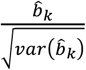 can be used to classify treatments according to their ‘distance’ from the average treatment.

Of note is that the above calculations assume normality of the estimated parameters 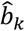. However, as 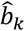 are not effect sizes expressed for example as logOR or mean differences, using them for hypothesis testing is not meaningful. Despite that, drawing 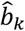 along with the associated 95% confidence intervals can be useful in capturing uncertainty around the ranking produced by relative treatment effects. Furthermore, by making some extra assumptions, 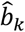 can be translated into absolute effects; for example, performing a meta-analysis of all treatment arms in the network can give an estimate of the expected outcome in the ‘average’ treatment. This estimate combined with 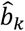 will then give absolute effects of each treatment.

### 3.1 Comparison of PreTAs with existing ranking metrics: theoretical considerations and empirical analysis

The, usually called, probability of being the best is a popular ranking metric which is interpreted as the probability of producing the best value in the outcome (pBV) in a network of interventions (e.g. large effects for a beneficial outcome, or small effects for a harmful outcome). While its derivation might be sensible in some cases, we should not overlook the fact that it only takes into account one tail of the treatment effects’ distributions; e.g. it does not account for the probability to produce a small effect on a beneficial outcome. SUCRAs and P-scores are useful summaries of the entire ranking distributions; suggested interpretations include *“the average proportion of competing treatments, which produce outcome values worse than treatment k”* and *“the mean extent of certainty that treatment k produces better values than all other treatments*” (7,19).

We performed an empirical comparison of the treatment hierarchies obtained with PreTA, pBV and SUCRA, calculated using parametric bootstrap in a frequentist framework. The agreement between ranking metrics was measured using Kendall’s tau. We used a previously described database of NMAs published until 2015 including networks of four or more interventions (1). We included networks with available outcome data in arm-level format, for which the primary outcome was analysed either as binary or as continuous. We used the effect measure used in the original review. Details about the inclusion criteria of the NMAs included in the database can be found in (1). The empirical analysis was performed with the use of the nmadb package in R (20).

In the following section, we illustrate our method in two networks of interventions, for which at least some disagreements between pBV, SUCRAs and PreTAs occur.

## 4 Worked examples

### 4.1 Network of antidepressants

We illustrate the derivation of the method using as an example a recently published NMA comparing the effectiveness of antidepressants for major depression (21). The primary efficacy outcome was response measured as 50% or greater reduction in the symptoms scales between baseline and 8 weeks of follow up and results were presented as ORs. The authors aimed at comparing active antidepressants and considered the inclusion of both head-to-head and placebo-controlled trials. The network comprised 522 double-blind, parallel, RCTs comparing 21 antidepressants or placebo. However, in line with previous empirical evidence (22,23), the authors have found evidence that the probability of receiving placebo decreases the overall response rate in a trial and dilutes differences between active compounds (24). Based on this ground, authors of this NMA (21) synthesized only head-to-head studies separately to estimate the relative efficacy of active interventions. Here, we will focus on the latter network that included 179 head-to-head studies comparing 18 antidepressants (Figure 1a).

**Figure 1.**
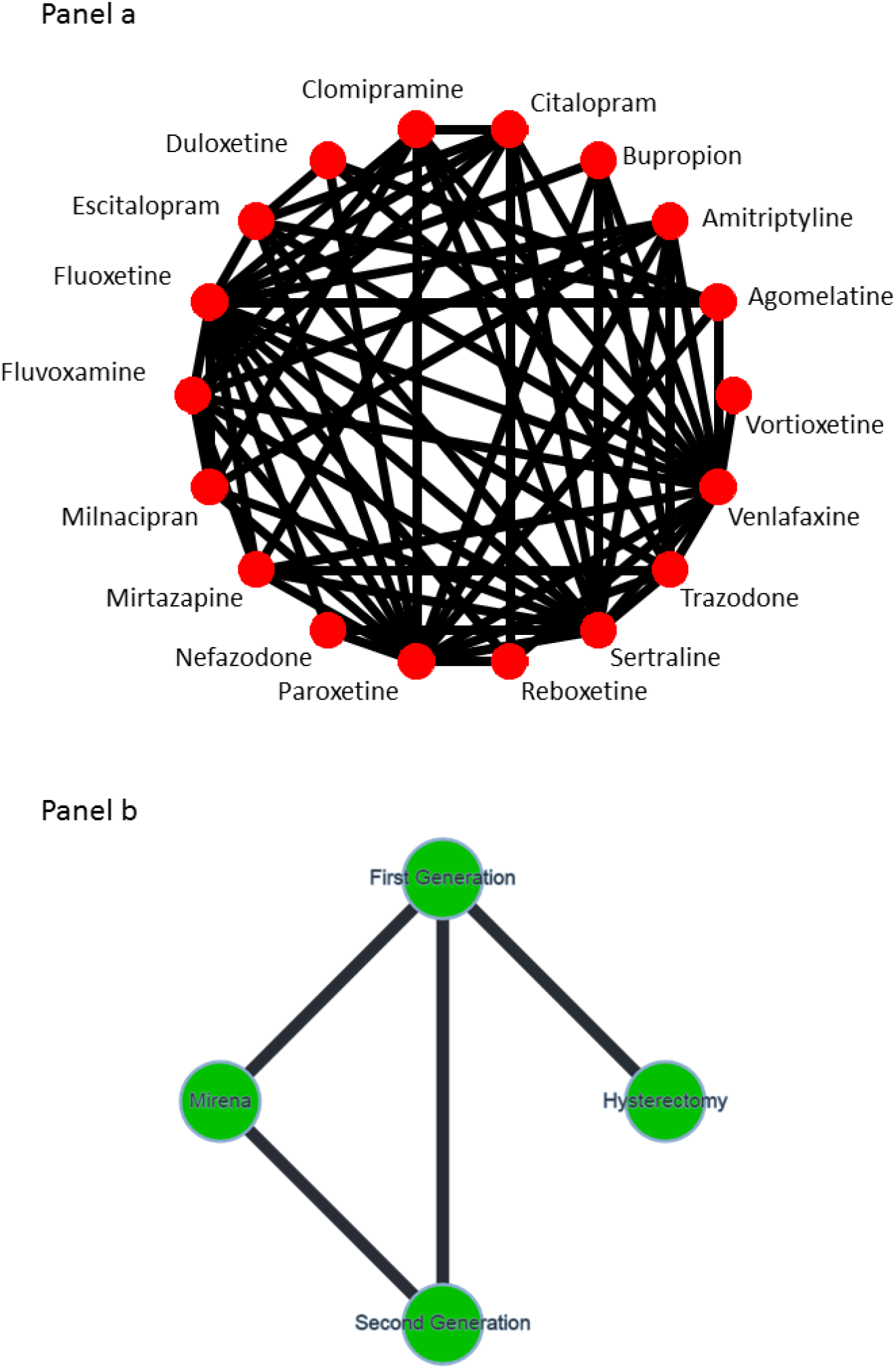
Panel a: Network plot of head-to-head randomized control trials comparing 18 antidepressants. Panel b: Network plot of head-to-head randomized control trials comparing 4 interventions for heavy menstrual bleeding. First and second generation interventions refer to endometrial destruction. Nodes and edges are unweighted.

Authors presented relative treatment effects between all pairs of the 18 antidepressants in a league table (figure 4 in (21)). When effect sizes are used to rank treatments, selecting a reference treatment against which to draw a forest plot of NMA effects is of particular importance. Although the choice of reference does not affect the estimates obtained, the uncertainty around NMA effects depends on the precision with which the selected reference treatment is associated. Figure 2 shows the relative treatment effects against fluoxetine and vortioxetine, the treatments that have been studied most and least respectively. While results are equivalent, choosing to present one over the other forest plot might implicitly lead to different interpretations on the similarity between the drugs based on visually inspecting the overlap of the confidence intervals.

**Figure 2.**
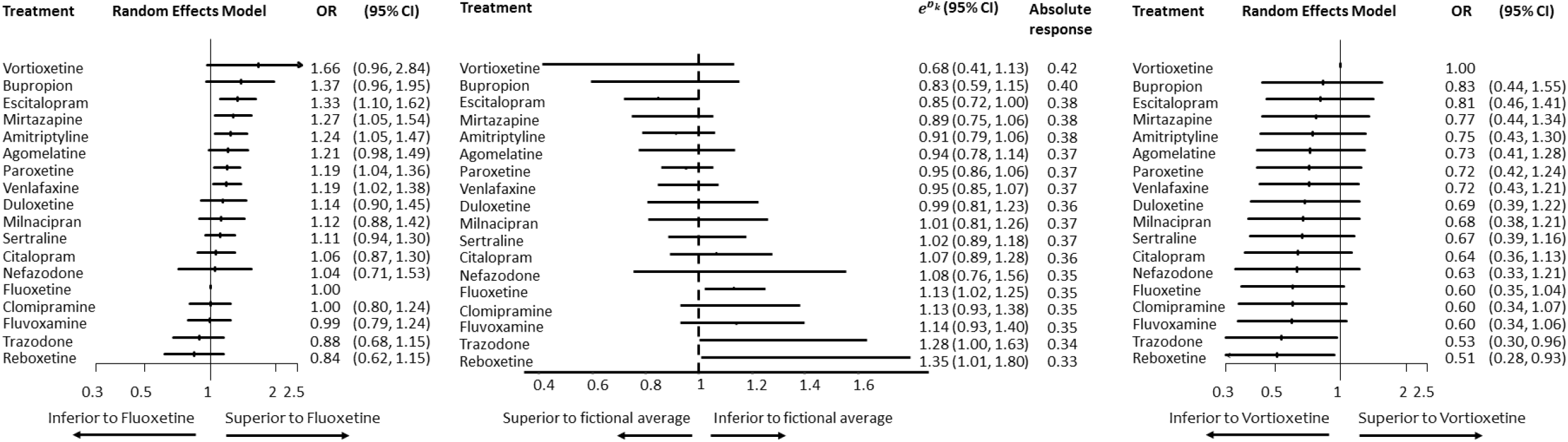
Odds ratios of each treatment versus fluoxetine, odds of each treatment versus odds of a fictional treatment of average response 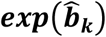 and odds ratios versus vortioxetine in the network of head-to-head studies comparing 18 antidepressants. OR: odds ratio; CI: confidence interval.

Figure 2 also shows the derived odds of each treatment versus the odds of a fictional treatment of average response with their confidence intervals. The line of no effect is included in the graph for illustration reasons, although 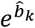 are not suited for hypothesis testing. The amount of uncertainty around the relative effects versus the average treatment is between the amount of uncertainty around the relative effects of fluoxetine and that of vortioxetine. In fact, presenting 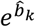 with their confidence intervals offers a solution to the ambiguity of selecting a reference treatment, in terms of the uncertainty around them and the consequent conclusions about similarity of treatments. Moreover, Figure 2 shows the approximated absolute responses for each treatment, assuming the expected risk of the average treatment calculated as the meta-analytic effect of all treatment responses.

Table 3 summarizes the ranking metrics for the network of antidepressants; pBV, the SUCRA and PreTAs are presented (6,25). Escitalopram, which is the first treatment according to PreTA, ranks second according to SUCRA and third according to pBV. The disagreement between PreTA and pBV is explained by the fact that pBV favours vortioxetine and bupropion over escitalopram because their effects are estimated with greater uncertainty. The small disagreement between PreTA and SUCRA reflects their different interpretations: vortioxetine, ranked first according to SUCRA, beats on average a larger proportion of treatments compared to escitalopram (0.90 versus 0.83) but escitalopram has a larger probability to be better than the fictional average treatment compared to vortioxetine (0.93 versus 0.87). Similarly, fluoxetine ranks last according to PreTA whereas it is followed by trazodone according to SUCRA.

**Table 3.** Ranking metrics for the network of antidepressants and ranks according to each ranking metric in parentheses. pBV: probability of producing the best value; SUCRA: surface under the cumulative ranking curve; PreTA: preferable than average.

|  | **pBV** | **SUCRA** | **PreTA** |
| --- | --- | --- | --- |
| Agomelatine | 0.01 (6) | 0.64 (6) | 0.74 (8) |
| Amitriptyline | 0.01 (7) | 0.71 (5) | 0.88 (4) |
| Bupropion | 0.20 (2) | 0.80 (3) | 0.87 (5) |
| Citalopram | 0.00 (17.5) | 0.37 (13) | 0.24 (13) |
| Clomipramine | 0.00 (15) | 0.26 (14) | 0.10 (14.5) |
| Duloxetine | 0.01 (9) | 0.52 (9) | 0.52 (9) |
| Escitalopram | 0.07 (3) | 0.83 (2) | 0.97 (1) |
| Fluoxetine | 0.00 (17.5) | 0.23 (16) | 0.01 (18) |
| Fluvoxamine | 0.00 (12.5) | 0.25 (15) | 0.10 (14.5) |
| Milnacipran | 0.01 (8) | 0.48 (10) | 0.46 (10) |
| Mirtazapine | 0.03 (4) | 0.75 (4) | 0.91 (3) |
| Nefazodone | 0.02 (5) | 0.38 (12) | 0.33 (12) |
| Paroxetine | 0.00 (10) | 0.62 (7) | 0.82 (6) |
| Reboxetine | 0.00 (15) | 0.09 (18) | 0.02 (16.5) |
| Sertraline | 0.00 (11) | 0.46 (11) | 0.38 (11) |
| Trazodone | 0.00 (15) | 0.12 (17) | 0.02 (16.5) |
| Venlafaxine | 0.00 (12.5) | 0.61 (8) | 0.78 (7) |
| Vortioxetine | 0.64 (1) | 0.90 (1) | 0.93 (2) |

Figure 3 shows the PreTAs for the 18 antidepressants; treatments around 0.5 are the treatments closest to the average treatment. Vortioxetine has the largest point estimate against the average treatment but its estimation comes with great uncertainty. Escitalopram versus average is more precisely estimated in favor of escitalopram and it is associated with the greatest PreTA (97%). Duloxetine and milnacipran are the treatments closest to the average treatment. The point estimate of nefazodone versus the average treatment is slightly larger than that of duloxetine. Due to the associated uncertainty, however, there is 34% probability that nefazodone is superior to the fictional average treatment, compared to 52% of duloxetine. Fluoxetine, clomipramine, fluvoxamine, trazodone and reboxetine are among the worst treatments in the network, either because of their point estimates against the average treatment or because of the respective precision in the estimation.

**Figure 3.**
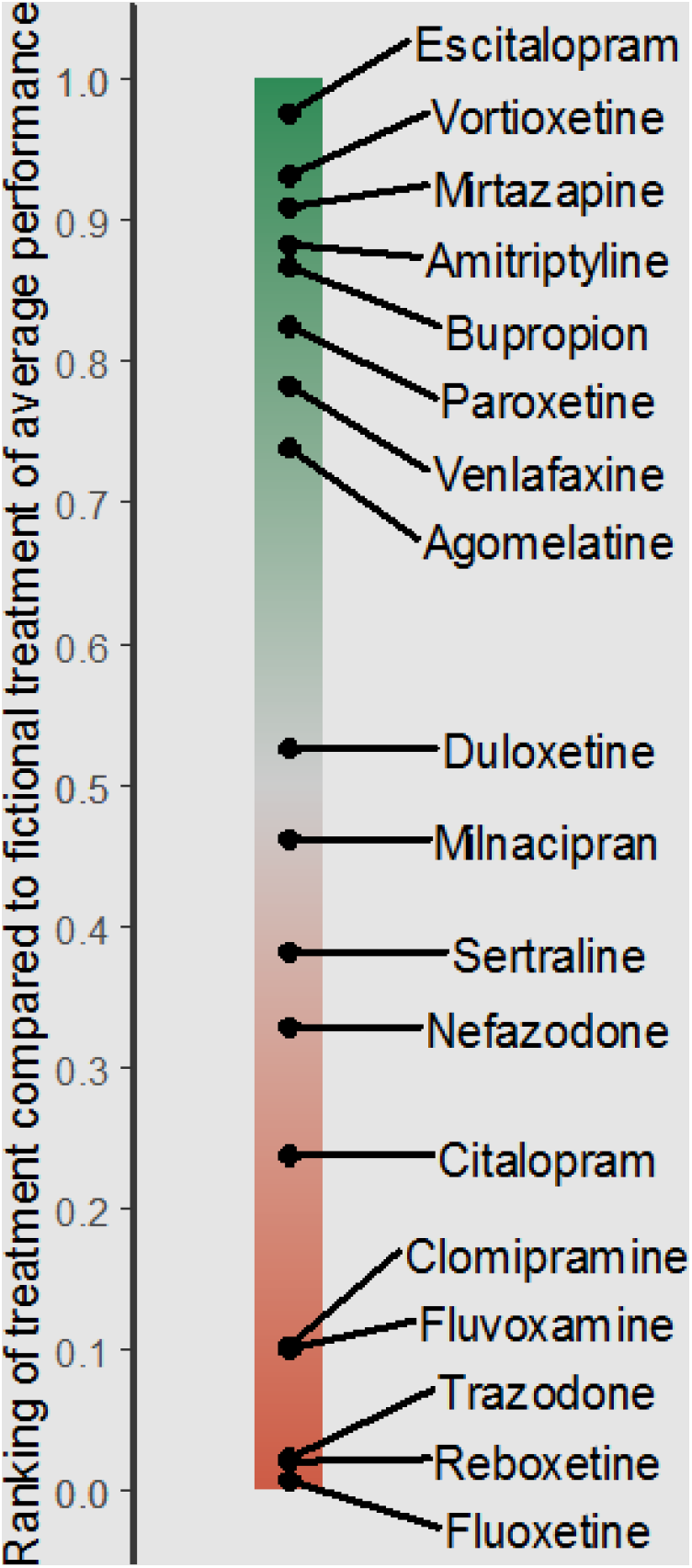
Classifier of interventions for the network of 18 antidepressants according to the probability of being preferable than average (PreTA).

### 4.2 Network of interventions for heavy menstrual bleeding

We use as a second example a network of interventions for the treatment of heavy menstrual bleeding. The following four interventions were compared: levenorgestel-releasing intrauterine system (Mirena), first generation endometrial destruction, second generation endometrial destruction and hysterectomy (26). The primary outcome was patients’ dissatisfaction at 12 months and the network included 20 studies (Figure 1b).

Figure 4 shows the treatment effects of the four treatments compared to a fictional average treatment and Appendix Figure 1 illustrates the relative position of each treatment according to its probability of being superior (green) or inferior (red) than the average treatment. There is a clear advantage of hysterectomy compared to the other three treatments with no treatment lying close to the ‘average treatment area’ (0.5 of PreTA).

**Figure 4.**
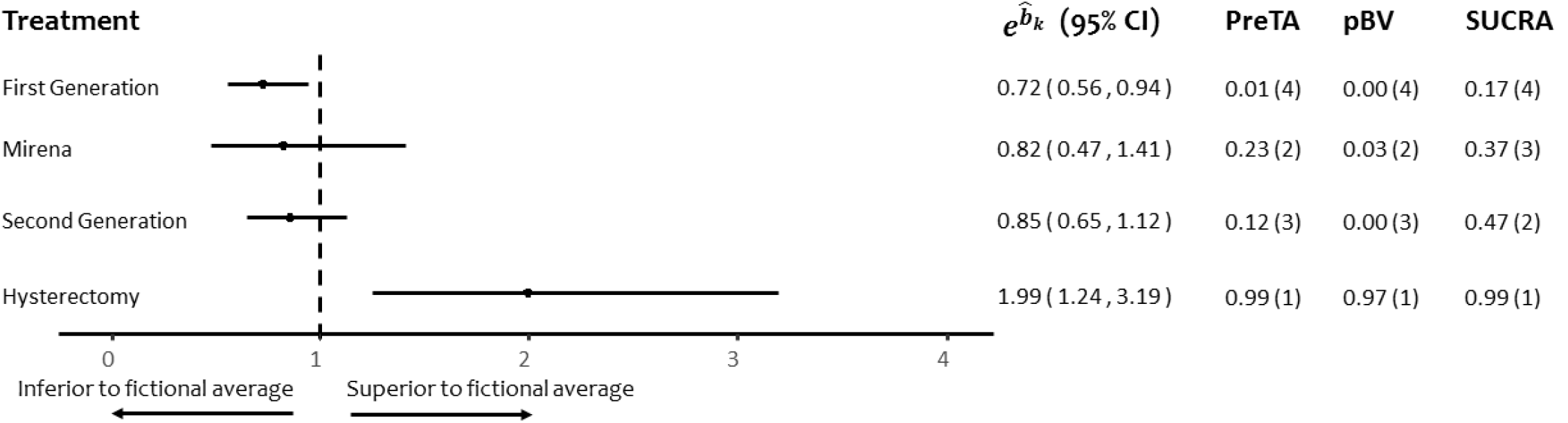
Odds of each treatment versus odds of a fictional treatment of average response 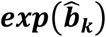, probability of each treatment being better than the average (PreTA), probability of producing the best value (pBV) and SUCRA in the network of head-to-head studies comparing 4 interventions for heavy menstrual bleeding. Numbers in parentheses under PreTA, pBV and SUCRA represent ranks. CI: confidence interval; PreTA: preferable than average; pBV: probability of producing the best value; SUCRA: surface under the cumulative ranking curve.

In this example, hysterectomy outperforms the other three treatments and ranks first according to all ranking metrics. Similarly, all ranking metrics agree that first generation endometrial destruction is the least preferable option (Figure 4). The disagreement between ranking metrics occurs for the second and third position between Mirena and second generation endometrial destruction. The two interventions are similar according to the point estimates but second generation is more precise. This leads to a greater certainty that second generation is worse than the average treatment compared to Mirena, resulting in a smaller PreTA. However, second generation beats on average more treatments than Mirena does since the relative effect of second generation is larger than that of Mirena; this results in a larger SUCRA for second generation than for Mirena.

## 5 Results of the empirical analysis

We ended up with 232 networks to be included in the empirical analysis. There was strong agreement between hierarchies obtained by PreTAs and SUCRAs, shown by a median Kendall’s tau (in the following called ‘correlation’) of 0.94 with interquartile range (IQR) 0.86 to 1.00). Almost half of the networks (101, 44%) had correlation of 1 while only two networks (1%) had correlation less than 0.6. The network with the smallest correlation (0.4) is shown in Appendix Figure 2 (27); while the hierarchy itself does not change much, PreTA and SUCRA disagree in proximity of treatments with similar point estimates and different precision. The agreement between PreTAs and pBV was lower with a median correlation of 0.74 (IQR 0.61 to 0.89) and 49 networks (21%) having correlation less than 0.6 (Appendix Figure 3).

As with all ranking metrics, any disagreements between PreTAs and pBV or SUCRAs are attributed to the different ways they incorporate uncertainty in the estimation. pBV favors treatments associated with uncertainty, as the tail of the distribution of treatments with uncertain effects is larger compared to the tail of the distribution for treatments with similar point estimate but high precision. The probability *P*_*kl*_ tends to 0.5 with increased 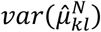; consequently, the greater the uncertainty associated with a treatment, the more its P-score tends to 0.5. A research paper describing theoretically the interpretation and the role of uncertainty in the various ranking metrics, as well as a detailed empirical analysis are in preparation (19,28).

## 6 Discussion

In this paper, we developed a new ranking metric, PreTA, interpreted as the probability of each treatment being preferable than a fictional treatment of average performance. The notion of the average treatment refers to the average absolute efficacy among the treatments included in the systematic review. Thus, as with all ranking metrics, the interpretation of PreTAs is subject to the set of treatments compared. PreTAs can be produced in all NMAs as long as the eligibility of treatments is well justified. The usefulness of the interpretation of the 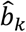 coefficients, however, depends on whether the the notion of an ‘average’ treatment makes sense.

In the presence of a reference treatment, e.g. placebo, a simple and intuitive non-probabilistic ranking metric can be obtained by ranking all relative effects against placebo. Authors of NMA often present estimated treatment effects against placebo or standard care in a forest plot, providing implicitly or explicitly a treatment hierarchy. While such a hierarchy might be appropriate in many settings, they assume that treatment effects against placebo are of primary interest for the analysis. This might not be the case in other healthcare areas where one or more established therapies exists (29) or where researchers are concerned about the quality of the evidence from placebo-controlled studies (30–32) and choose to, exclusively or complementary, analyse a network without placebo. Moreover, it should be taken into account that the amount of data associated with the reference treatment might have an impact on the judgement regarding the similarity of the treatments, when such a judgement is made by visually inspecting a forest plot of NMA effects. Point estimates against the fictional average treatment provide a solution to this ambiguity. Furthermore, data from registries can be assumed to approximate the response of an average treatment, as participants may take any of the available interventions. Thus, using such external data, absolute effects can be approximated using the point estimates against the average.

Alternative methods to avoid the reference group coding have been suggested in the literature. The application of quasi-variances (33), independently proposed as ‘floating absolute risks’ in epidemiology (34), do avoid setting a reference group. However, the scope of their use pertains to approximating a set of variances of the model contrasts such that the variances between any linear combination of contrasts can be derived without the disposal of the covariance matrix (35). Thus, quasi-variances approaches target a different problem from the model described in this paper and the relevance of the estimated quantities to NMA is not clear.

Producing a treatment hierarchy in NMA is popular, with 43% of published NMAs presenting at least one ranking metric (1), but also debatable. Recent developments tackle common criticisms against ranking metrics, pertaining to arguments that they are unstable (36,37), uncertain (38), do not differentiate between clinically important and unimportant differences (4,39), do not account for multiple outcomes (40) and are not accompanied by a measure of uncertainty (41). In particular, recent developments include extensions of P-scores for two or more outcomes (42), incorporation of clinically important values in their calculation (42), application of multiple-criteria decision analysis (43) and partial ordering of interventions according to multiple outcomes (44). PreTAs can be easily extended to incorporate clinically important values as shown in (42); such probabilities will then be interpreted as the probability of a treatment being better than the average by at least a certain value.

PreTA is a viable alternative to existing ranking metrics, that can be interpreted as a probability and takes into account the entire ranking distribution. As it is also the case with PreTA, all existing ranking metrics use the distribution of NMA treatment effects to produce a hierarchy of the treatments. This hierarchy can be based either on probabilities like “which is the probability that each treatment produces the best outcome value” or “which is the probability of treatment A beating treatment B” or summaries of these probabilities. Rankograms visualise the entire ranking distributions for each treatment and SUCRAs, P-scores and mean ranks summarise these probabilities in a single number for each treatment. The interpretation of these summaries is, however, not always straightforward. The development of PreTAs enriches the decision-making arsenal with a presentational and ranking tool, which can be interpreted in a clinically meaningful way.

## Data Availability

Outcome data and the code for applying our methods are available in https://github.com/esm-ispm-unibe-ch/alternativenma.

## 7 Declarations

## Conflicts of interest

TAF reports personal fees from Mitsubishi-Tanabe, MSD and Shionogi and a grant from Mitsubishi-Tanabe, outside the submitted work; TAF has a patent 2018-177688 pending.

## Funding

AN, VC, TP and GS were supported by project funding (Grant No. 179158) from the Swiss National Science Foundation.

## Authors’ contributions

AN conceived the idea, contributed to the modelling, produced the results and wrote the R code and the first draft of the manuscript. VC contributed to the analysis. TP contributed to the modelling and to the R code. DM, TAF and GS contributed to the modelling, reviewed the R code and contributed to the writing. All authors read and approved the final manuscript.

## 8 Figure legends

**Appendix Figure 1.**
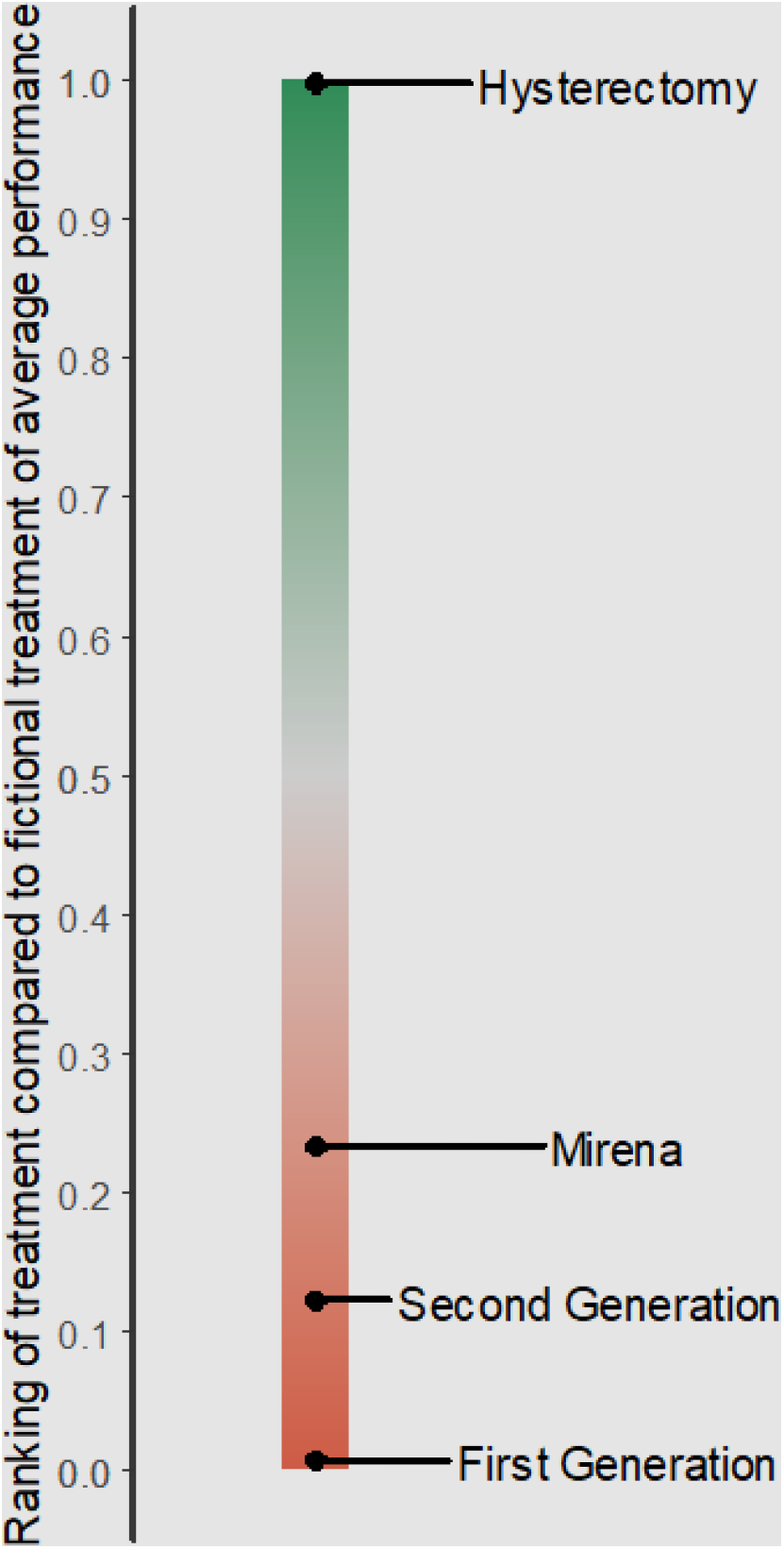
Classifier of interventions for the network of four interventions for heavy menstrual bleeding according to the probability of being preferable than average (PreTA).

**Appendix Figure 2.**
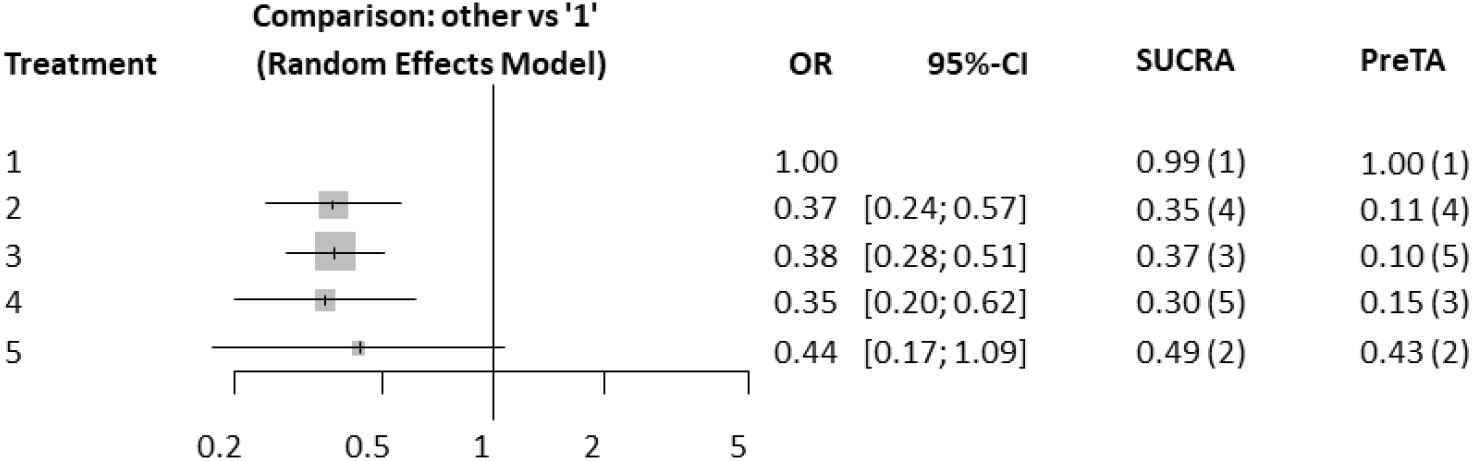
Odds ratios, probability of each treatment being better than the average (PreTA) and SUCRA in the network with the smallest correlation between PreTA and SUCRA. Numbers in parentheses under PreTA, pBV and SUCRA represent ranks. OR: odds ratio; CI: confidence interval; PreTA: preferable than average; pBV: probability of producing the best value; SUCRA: surface under the cumulative ranking curve.

**Appendix Figure 3.**
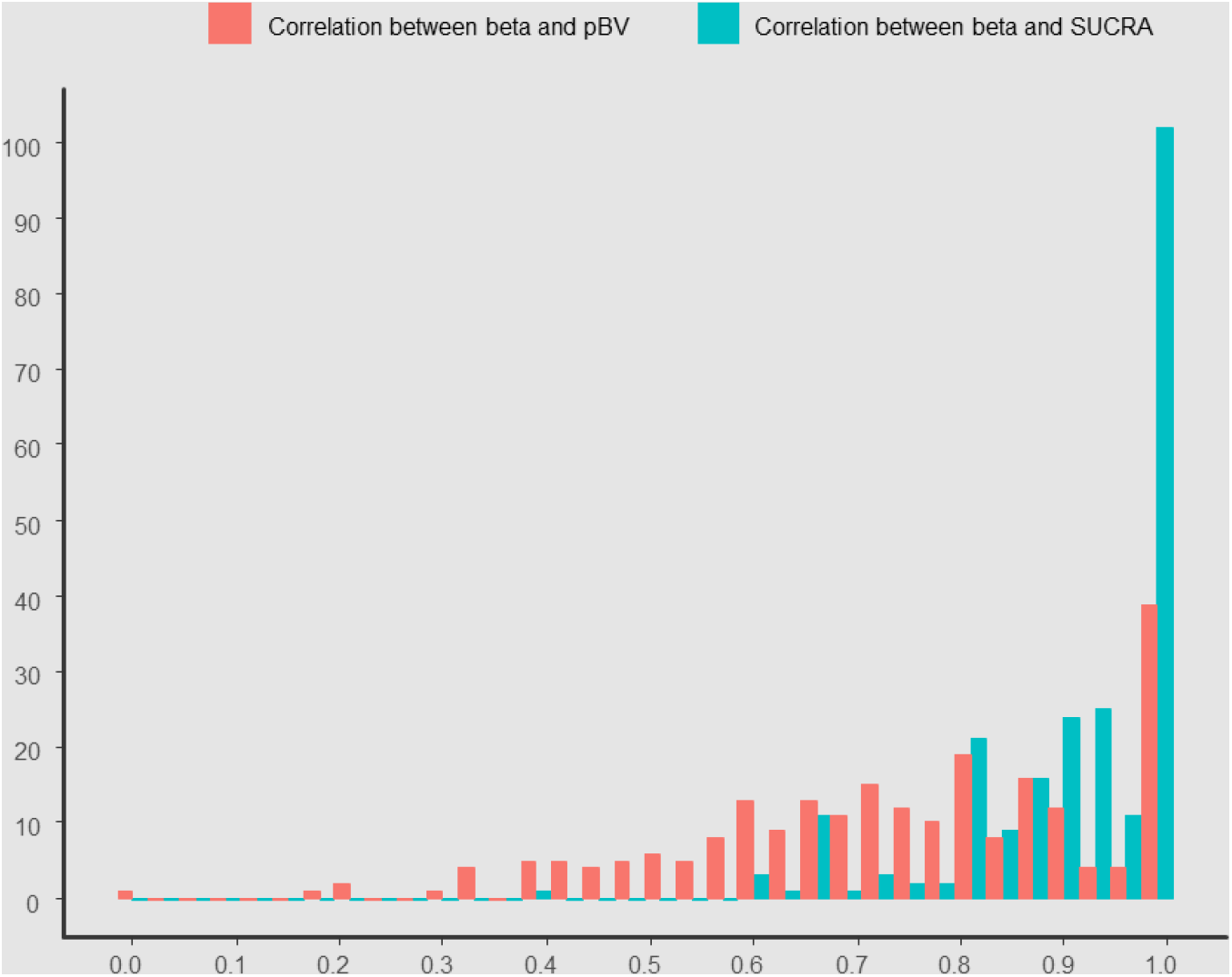
Histogram of correlation measured as Kendall’s tau between probability of being better than average (PreTA) and probability of producing the best value (red) and between PreTA and surface under the cumulative ranking curve (SUCRA) (blue). pBV: probability of producing the best value; SUCRA: surface under the cumulative ranking curve; PreTA: preferable than average.

## 9 Highlights

**What is already known:** A treatment hierarchy is often of interest to users of network meta-analysis. However, interpretation of ranking metrics is often challenging.

**What is new:** We present a new ranking metric (PreTA: Preferable Than Average) interpreted as the probability that a treatment is better than a fictional treatment of average performance.

**Potential impact for Review Synthesis Methods readers outside the authors’ field:** The proposed ranking metric uses the entire ranking distribution and it is interpreted as a probability. It can be used as a viable alternative to existing ranking metrics in systematic reviews with multiple interventions.

